# Association between rRT-PCR test results upon admission and outcome in hospitalized chest CT-Positive COVID-19 patients; a provincial retrospective cohort with active follow-up

**DOI:** 10.1101/2020.04.21.20074641

**Authors:** Saeed Nemati, Hamid Reza Najari, Anita Eftekharzadeh, Amir Mohammad Kazemifar, Ali Qandian, Pedram Fattahi, Maedeh Zokaei Nikoo, Shiva Leghaei, Mohammad Reza Rouhollahi

## Abstract

**Background:** The Covid-19 pandemic imposed the most devastating challenge on healthcare systems worldwide. Iran was among the first countries that had to confront serious shortages in RT-PCR testing for SARS-CoV-2 and ventilators availabilities throughout the COVID-19 outbreak. This study aimed to investigate the clinical course of hospitalized COVID-19 patients with different rRT-PCR test results during the first 3 weeks of the outbreak in Qazvin province, Iran.

**Methods:** For this retrospective cohort study, data of hospitalized patients primarily diagnosed as having COVID-19 in all 12 centers across the whole Qazvin province during Feb 20-Mar 11, 2020 was analyzed. A multivariate logistic regression model was applied to assess the independent associates of death among COVID-19 patients.

**Results:** 998 patients (57% male, median age 54 years) with positive chest CT-scan changes were included in this study. Among them, 558 patients were examined with rRT-PCR test and 73·8% tested positive. Case fatality rate was 20·68% and 7·53% among test-positive and test negative hospitalized patients, respectively. While only 5·2% of patients were ICU admitted, case fatality rates outside ICU were 17·70% and 4·65% in test-positive and test-negative non-ICU admitted patients, correspondingly. The independent associates of death were age ≥ 70 years, testing positive with rRT-PCR test, having immunodeficiency disorders and ICU admission.

**Conclusions:** Hospitalized COVID-19 patients with mild symptoms despite positive chest CT changes and major comorbidities were more probable to have negative rRT-PCR test result, hence lower case fatality rate and a more favorable outcome.

## Introduction

The first report of COVID-19 in Iran was officially announced on February 19, 2020, from the city of Qom in central Iran.^(1)^ Shortly thereafter, cases of infection with the novel coronavirus were reported from all over the country. The province of Qazvin, including its capital - city of Qazvin which is located 200 km away from Qom, the coronavirus epicenter - was affected almost early during the COVID-19 epidemic in Iran. The province had a population of 1,300,000 people and accounted for 8% of the country’s economy.^(2)^ The first patient with a proven COVID-19 diagnosis in the province – a man in his mid-80s - was announced dead on March 1. This was also the first related death that was reported in Qazvin province.

To address the encountered challenges, the first edition of Iran National Interim Guidance on COVID-19 Diagnosis and Treatment (INIGCDT), which was published on February 24, limited the indications for rRT-PCR testing to detect SARS-CoV-2 to those admitted with severe respiratory signs and symptoms or admitted patients with fever whose chest imaging revealed pulmonary infiltration.^(3)^

During the less known COVID-19 outbreak in Iran, it was presumed that the rapid progression of infected and thus hospital admitted patients would soon overload the medical system. The problem would be even more prominent at the provincial level. In the “pre-overload” phase of the novel coronavirus epidemic in Qazvin - just before the rapid hospitalization rate across the province reached its peak capacity - a unique opportunity was provided to assess not only the clinical pattern and outcome of the disease, but also to evaluate the performance of care centers throughout the province. The results of such an assessment can further be employed as a basis for more precise admission criteria aimed at saving more lives in the subsequent critical phases of the epidemic.

In light of the above issues and considering that since the emergence of the novel coronavirus epidemic in Iran, little has been documented about the course and outcome of the affected patients in the country, this retrospective cohort study aimed to investigate the clinical pattern and outcome of patients admitted to the hospitals and intensive care facilities of Qazvin province, Iran in a critical 3-weeks period - from February 20 to March 11, 2020 - with a primary diagnosis of COVID-19 but different rRT-PCR test results.

## Methods

In this retrospective cohort study, patients who were admitted to all 12 hospitals across Qazvin province, Iran, during February 20 - March 11 with a primary diagnosis of COVID-19 (see definition) were included and followed-up until March 27, 2020.

Upon the outbreak, the Qazvin University School of Medicine – as the provincial health governor - provided an electronic data entry platform to collect epidemiologic and clinical data related to patients with COVID-19 and mandated all the involved hospitals within the province to employ the database. Data was collected from patients’ medical records and entered into the electronic database during the patients’ admission period. This included epidemiologic and clinical data, and patients’ exposure history as well as their results of COVID-19 rRT-PCR test and the clinical outcome (discharge, death). The patients who were admitted to hospital during March 5 - March 11, 2020 (the last week of this study) were later followed-up by phone to confirm their outcome.

This study was approved by the Research Ethic Committee of Qazvin University of Medical Sciences (QUMS-EC).

### Criteria and definitions

Patients were admitted with a primary diagnosis of COVID-19 based on INIGCDT.^(3)^ According to the guidance, patients were admitted with a primary diagnosis of COVID-19 if they had symptoms of fever, cough or myalgia (referred here as minor clinical criteria) and A) presented with either respiratory distress, low pulse oximetry reading (Sp02 < 93%), respiratory rate (RR) > 30 (addressed here as major clinical criteria) or had decreased level of consciousness or B) they were among the high risk groups who had an underlying medical condition along with suggestive chest X-ray or CT scan changes for COVID-19 (see below). Further laboratory and chest imaging studies were made during the patients’ admission course.

A definitive diagnosis of COVID-19 was made by rRT-PCR test to detect SARS-CoV-2 in patients’ respiratory secretions. The test became available through Iran Pasteur Institute in early February, 2020. Throat-swab and lower respiratory secretion (in intubated patients) specimens were obtained from patients and sent for SARS-CoV-2 PCR test during their admission course. However, due to a serious shortage in the test availability in Iran at that time, no re-examination was made for the patients.

Fever was defined as oral temperature > 37·8ºC. Shortness of breath and decreased level of consciousness were defined upon triage physicians’ clinical judgement. Underlying medical conditions that justified patients’ admission as proposed by INIGCDT included a history of cardiovascular diseases, pulmonary diseases, diabetes, hypertension, cancer, HIV infection, organ transplantation, Chest X-ray or CT scan changes suggestive for COVID-19 consisted of bilateral patchy infiltration with rapid progression toward ground glass opacity (GGO).^(3)^

Patients were admitted to intensive care units if they had persistent hypoxemia, decreased level of consciousness, hemodynamic instability, or hypercapnia.

People were discharged from the hospital if they had no fever for more than 48-72 hours, had SpO2 > 93% while breathing room air, their respiratory and clinical symptoms and signs improved, and their serial chest imaging showed remarkable improvement.

### Patients follow-up

The patients who were admitted during the third week of this study period (March 5 - March 11, 2020) were actively followed-up using phone interviews on March 27. Among them, 436 patients (response rate=76·7%) were reached.

### Statistical analysis

Descriptive analysis was run using median (IQR). Patients were categorized based on weeks of admission, patients’ outcome inside and outside the ICU and their rRT-PCR testing status. Case fatality rate (CFR) were assessed in each group. The Chi-Square test was employed to assess differences in study variables among the above categories of patients.

A multiple logistic regression model was applied to identify associated factors of death in COVID-19 infected patients. Odds Ratio (OR) and 95% CI for each contributing factor. Statistical analysis was performed using the Stata statistical software package (StataCorp. 2014. Stata Statistical Software: Release 14.1, College Station, TX: StataCorp LP) and a P value less than 0·05 was considered as significant.

## Results

Data of 1197 patients who were admitted to Qazvin province hospitals was entered into the Qazvin province COVID-19 electronic database during Feb 20-March 11, 2020. Among them, a group of 191 PCR-negative patients who were admitted to non-COVID-19 wards was left out from analysis, however, their clinical course was followed up as well. The case fatality rate (CRF) among these patients was 3·1% and none of them were admitted to intensive care units. Furthermore, 8 patients whose death occurred within the first 24 hours of their admission were excluded from the study (Fig 1).

**Figure 1.**
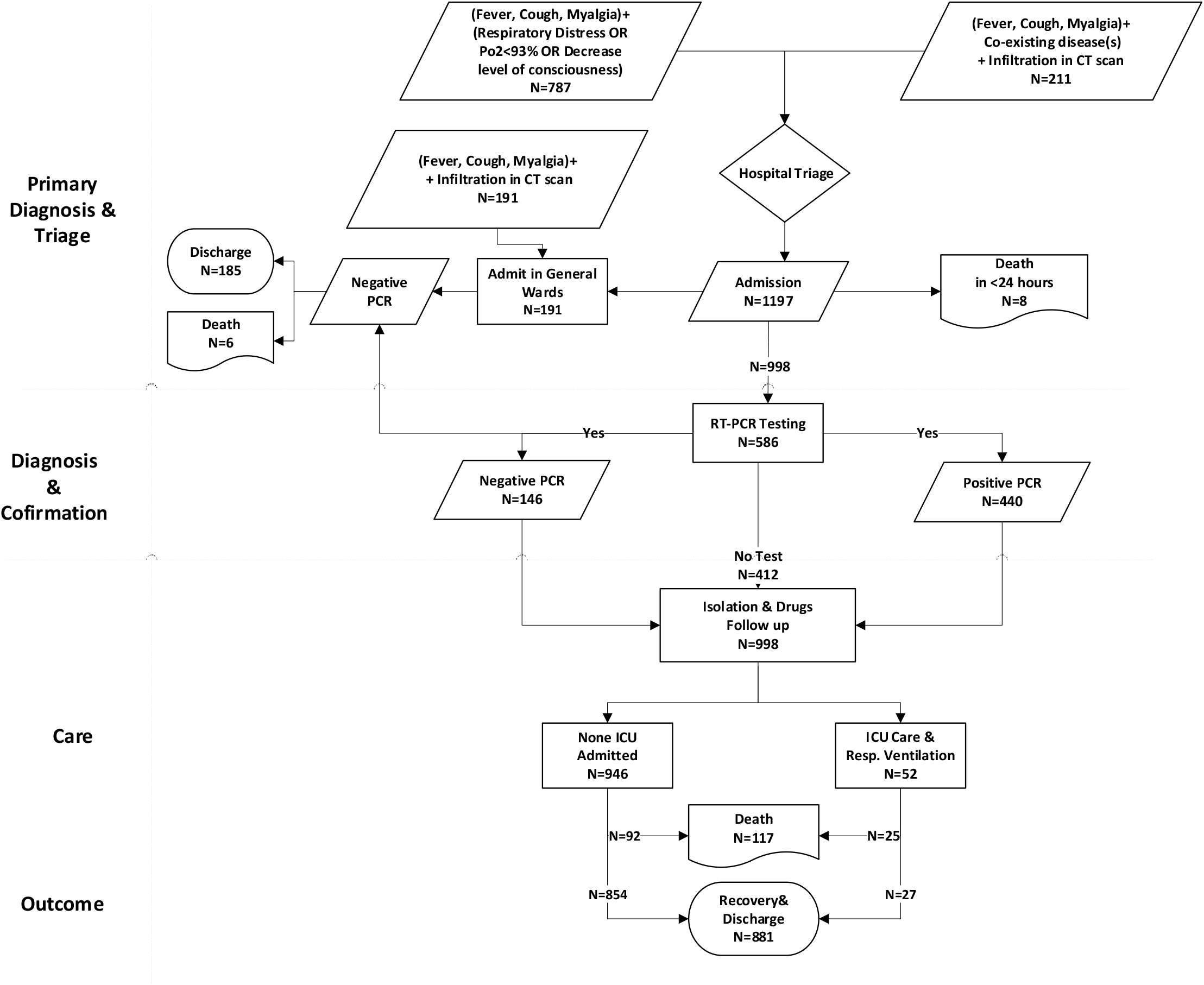
The study patients’ flow diagram

A total of 998 patients (57% male) with a median (IQR 25-75) age of 54(25) years were analyzed. From among them, 558 of the patients were tested for COVID-19 and 412 patients had positive results, while rRT-PCR test was negative in 146 cases. ICU care was provided for 5·2% patients, while others patients were isolated in the designated wards receiving low-flow oxygen and medical therapy. (Table 1)

**Table 1:**
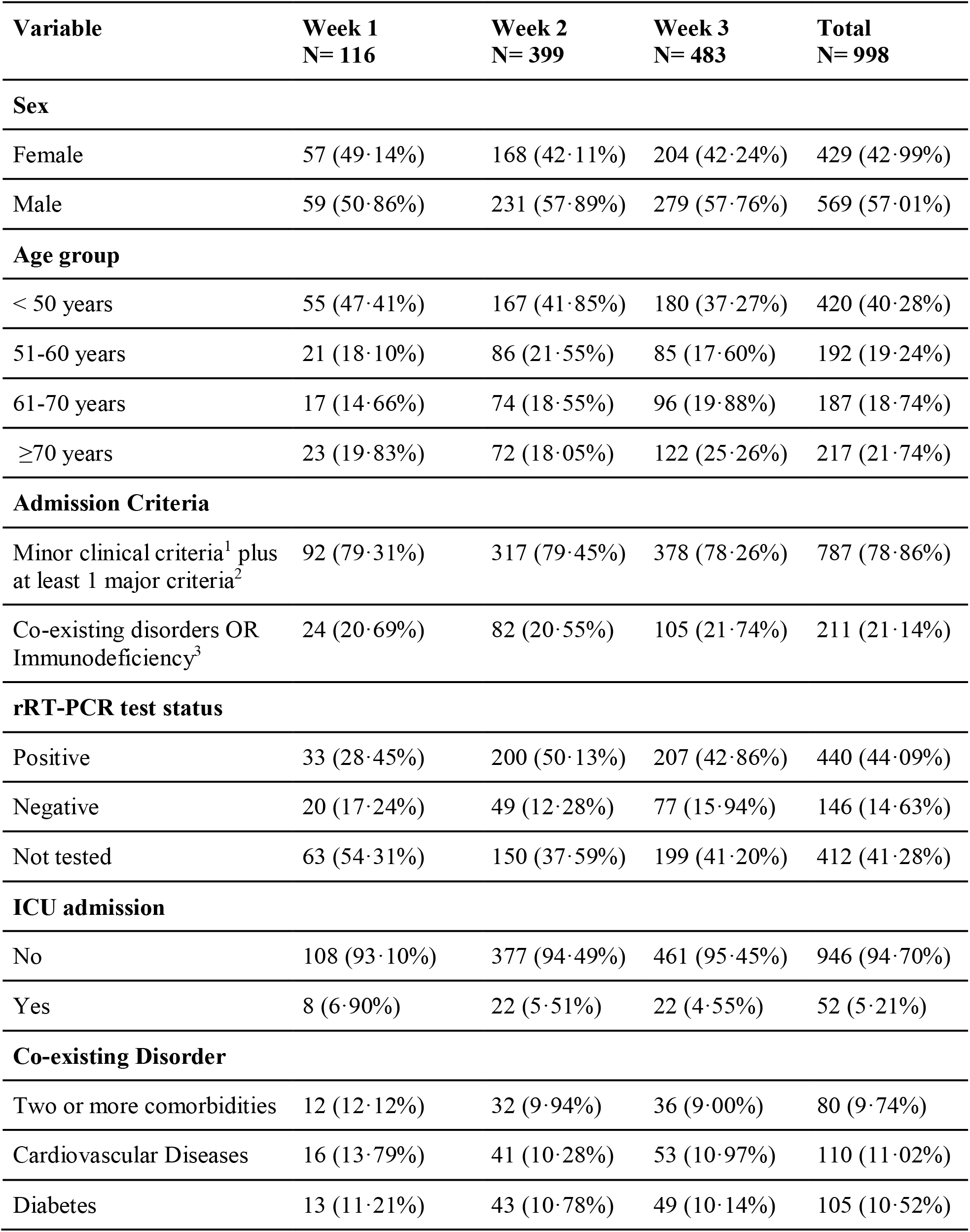
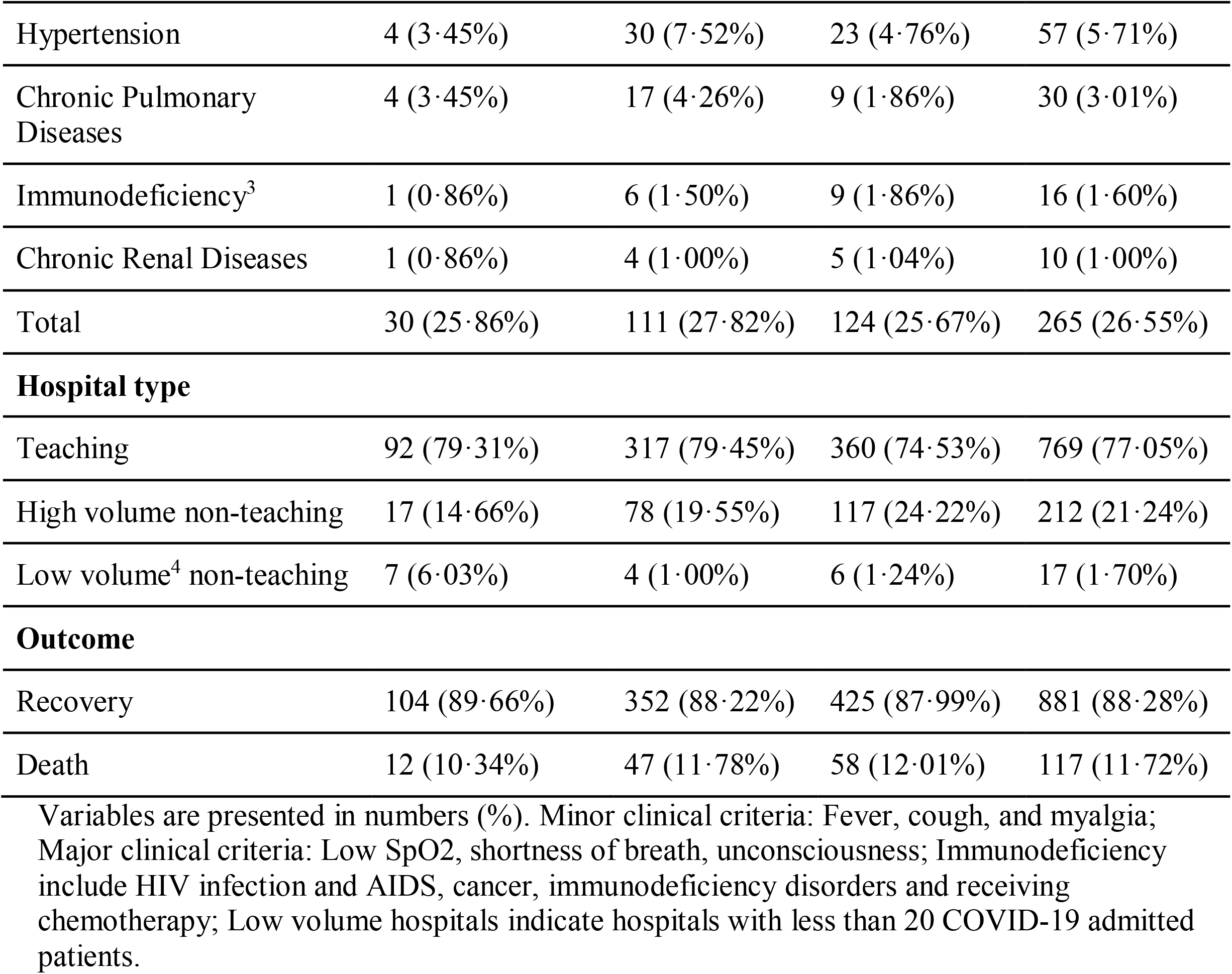
Basic characteristics of the study participants

| **Variable** | **Week 1**<br>**N= 116** | **Week 2**<br>**N= 399** | **Week 3**<br>**N= 483** | **Total**<br>**N= 998** |
| --- | --- | --- | --- | --- |
| **Sex** |  |  |  |  |
| Female | 57 (49·14%) | 168 (42·11%) | 204 (42·24%) | 429 (42·99%) |
| Male | 59 (50·86%) | 231 (57·89%) | 279 (57·76%) | 569 (57·01%) |
| **Age group** |  |  |  |  |
| < 50 years | 55 (47·41%) | 167 (41·85%) | 180 (37·27%) | 420 (40·28%) |
| 51-60 years | 21 (18·10%) | 86 (21·55%) | 85 (17·60%) | 192 (19·24%) |
| 61-70 years | 17 (14·66%) | 74 (18·55%) | 96 (19·88%) | 187 (18·74%) |
| ≥70 years | 23 (19·83%) | 72 (18·05%) | 122 (25·26%) | 217 (21·74%) |
| **Admission Criteria** |  |  |  |  |
| Minor clinical criteria<sup>1</sup> plus at least 1 major criteria<sup>2</sup> | 92 (79·31%) | 317 (79·45%) | 378 (78·26%) | 787 (78·86%) |
| Co-existing disorders OR Immunodeficiency<sup>3</sup> | 24 (20·69%) | 82 (20·55%) | 105 (21·74%) | 211 (21·14%) |
| **rRT-PCR test status** |  |  |  |  |
| Positive | 33 (28·45%) | 200 (50·13%) | 207 (42·86%) | 440 (44·09%) |
| Negative | 20 (17·24%) | 49 (12·28%) | 77 (15·94%) | 146 (14·63%) |
| Not tested | 63 (54·31%) | 150 (37·59%) | 199 (41·20%) | 412 (41·28%) |
| **ICU admission** |  |  |  |  |
| No | 108 (93·10%) | 377 (94·49%) | 461 (95·45%) | 946 (94·70%) |
| Yes | 8 (6·90%) | 22 (5·51%) | 22 (4·55%) | 52 (5·21%) |
| **Co-existing Disorder** |  |  |  |  |
| Two or more comorbidities | 12 (12·12%) | 32 (9·94%) | 36 (9·00%) | 80 (9·74%) |
| Cardiovascular Diseases | 16 (13·79%) | 41 (10·28%) | 53 (10·97%) | 110 (11·02%) |
| Diabetes | 13 (11·21%) | 43 (10·78%) | 49 (10·14%) | 105 (10·52%) |
| Hypertension | 4 (3·45%) | 30 (7·52%) | 23 (4·76%) | 57 (5·71%) |
| Chronic Pulmonary Diseases | 4 (3·45%) | 17 (4·26%) | 9 (1·86%) | 30 (3·01%) |
| Immunodeficiency<sup>3</sup> | 1 (0·86%) | 6 (1·50%) | 9 (1·86%) | 16 (1·60%) |
| Chronic Renal Diseases | 1 (0·86%) | 4 (1·00%) | 5 (1·04%) | 10 (1·00%) |
| Total | 30 (25·86%) | 111 (27·82%) | 124 (25·67%) | 265 (26·55%) |
| **Hospital type** |  |  |  |  |
| Teaching | 92 (79·31%) | 317 (79·45%) | 360 (74·53%) | 769 (77·05%) |
| High volume non-teaching | 17 (14·66%) | 78 (19·55%) | 117 (24·22%) | 212 (21·24%) |
| Low volume<sup>4</sup> non-teaching | 7 (6·03%) | 4 (1·00%) | 6 (1·24%) | 17 (1·70%) |
| **Outcome** |  |  |  |  |
| Recovery | 104 (89·66%) | 352 (88·22%) | 425 (87·99%) | 881 (88·28%) |
| Death | 12 (10·34%) | 47 (11·78%) | 58 (12·01%) | 117 (11·72%) |

Variables are presented in numbers (%). Minor clinical criteria: Fever, cough, and myalgia; Major clinical criteria: Low SpO<sub>2</sub>, shortness of breath, unconsciousness; Immunodeficiency include HIV infection and AIDS, cancer, immunodeficiency disorders and receiving chemotherapy; Low volume hospitals indicate hospitals with less than 20 COVID-19 admitted patients.

Comparing the study participants’ characteristics between the four groups of survivors and non-survivors outside and inside the ICU, no sex difference in CFR was found (p=0·674). Among non-survivors, except for the age group < 50 years, death rates were higher outside the ICU than inside intensive care units. Also, the difference was more prominent among patients aged>70 (23·04% vs 4·15%). Notably, in the rRT-PCR test-positive group, a 20·68% CFR was observed (Table 2).

**Table 2:**
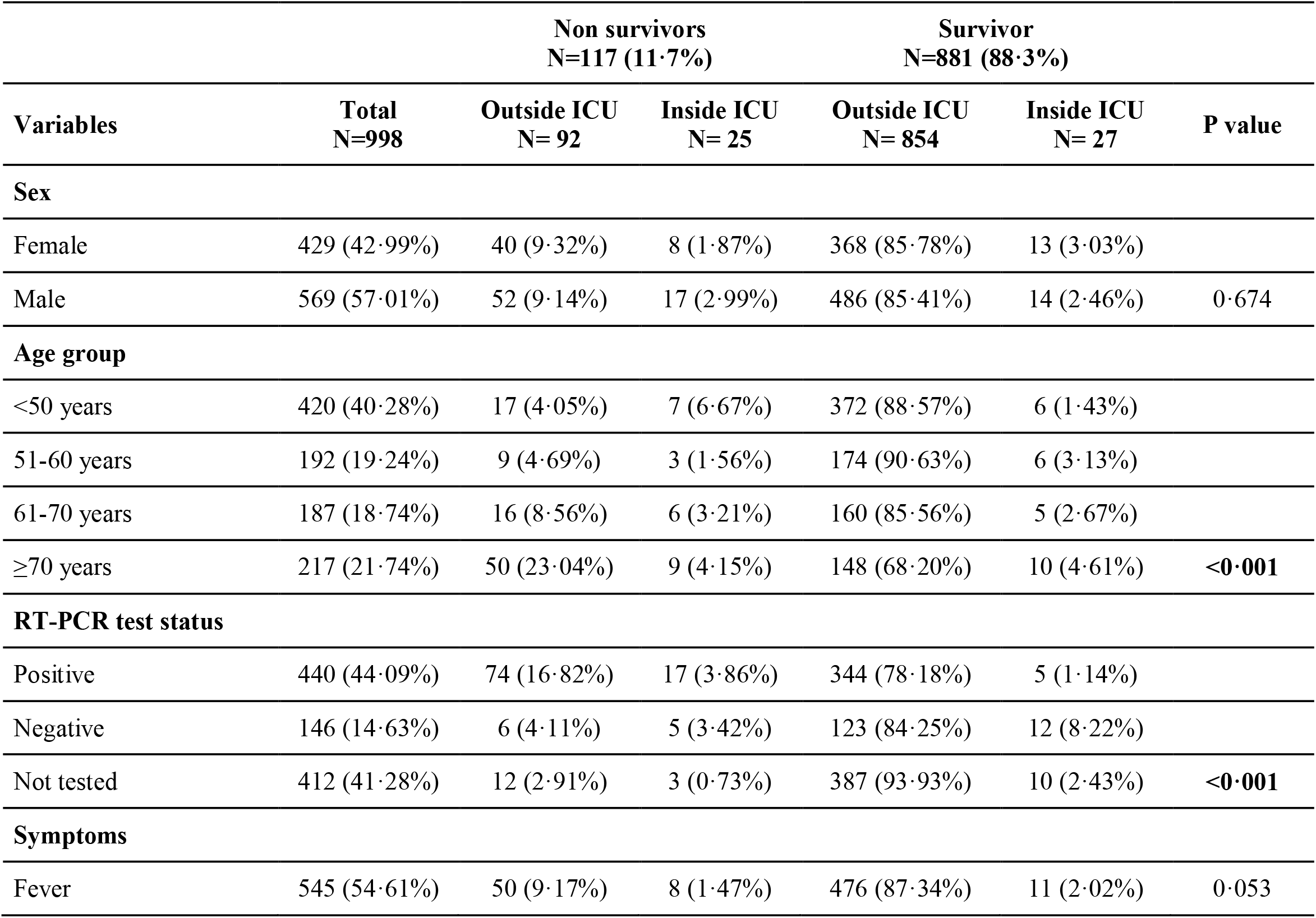
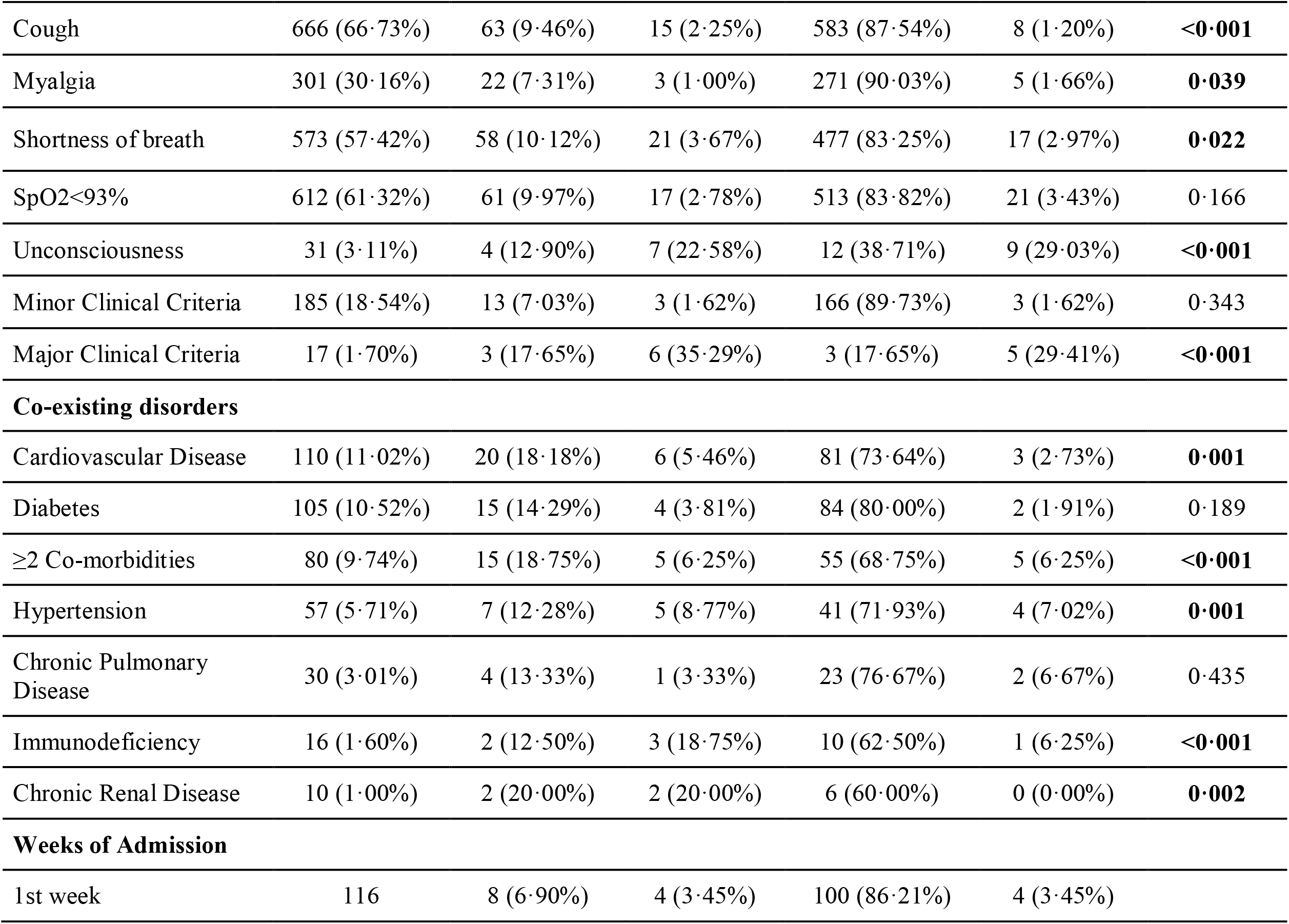
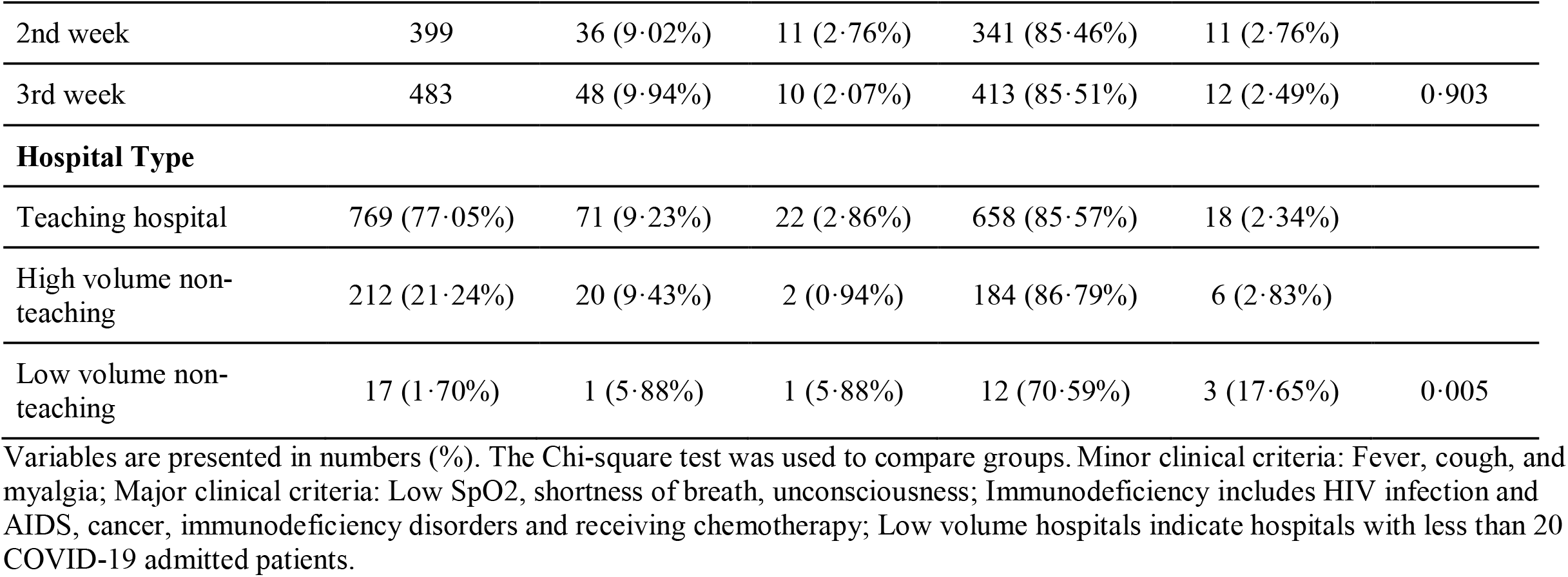
Comparing different prognostic factors between survivors and non-survivors in ICU admitted and non-ICU admitted patients

When comparing the study participants based on their PCR test results, it was found that while individuals aged > 70 years were more tested than the younger age groups, there was no statistically significant age difference between positive and negative rRT-PCR patients. Moreover, patients receiving ICU care were more tested, while in non-ICU admitted patients, positive results were more prevalent. (Table 3).

**Table 3:**
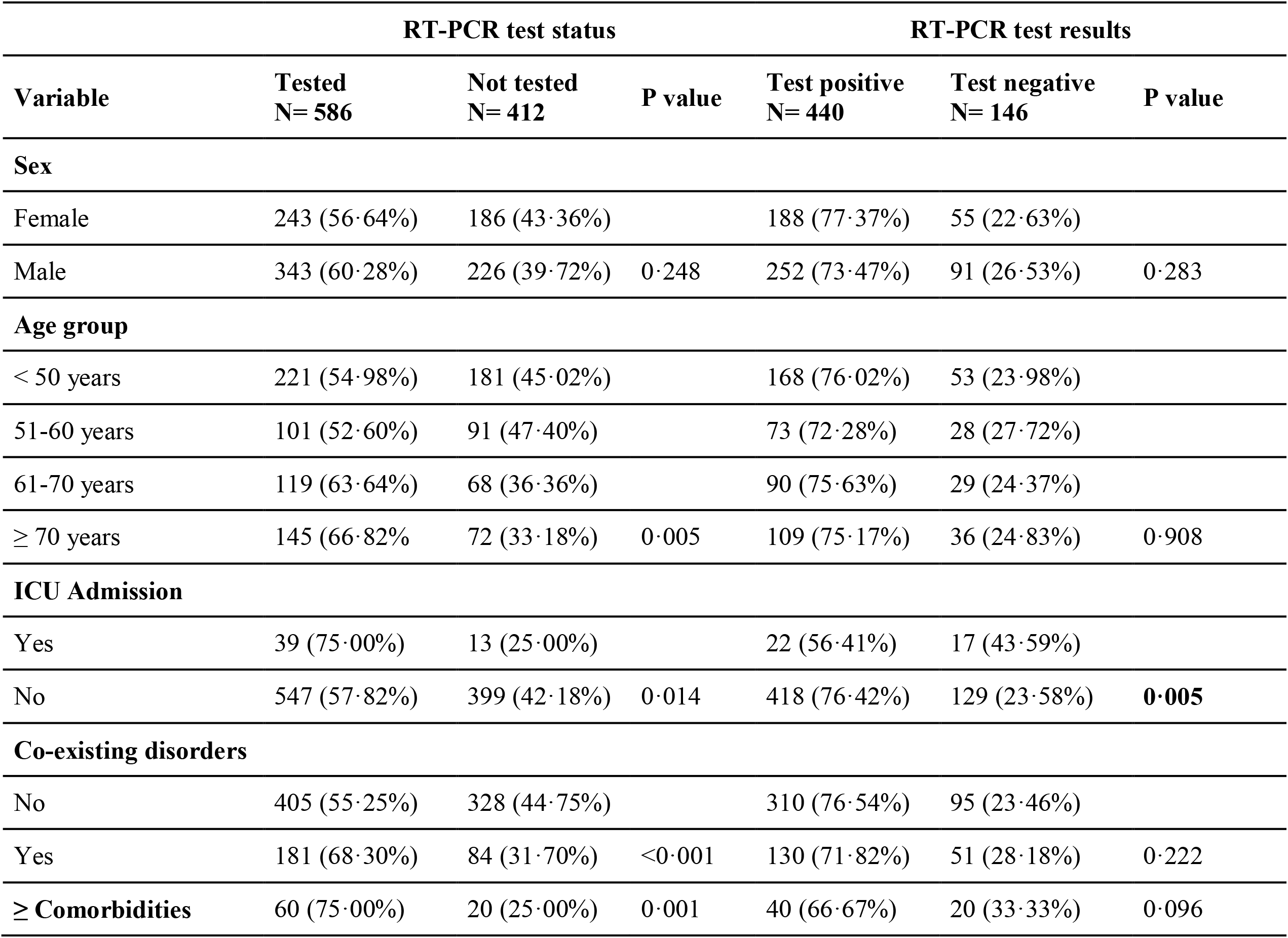
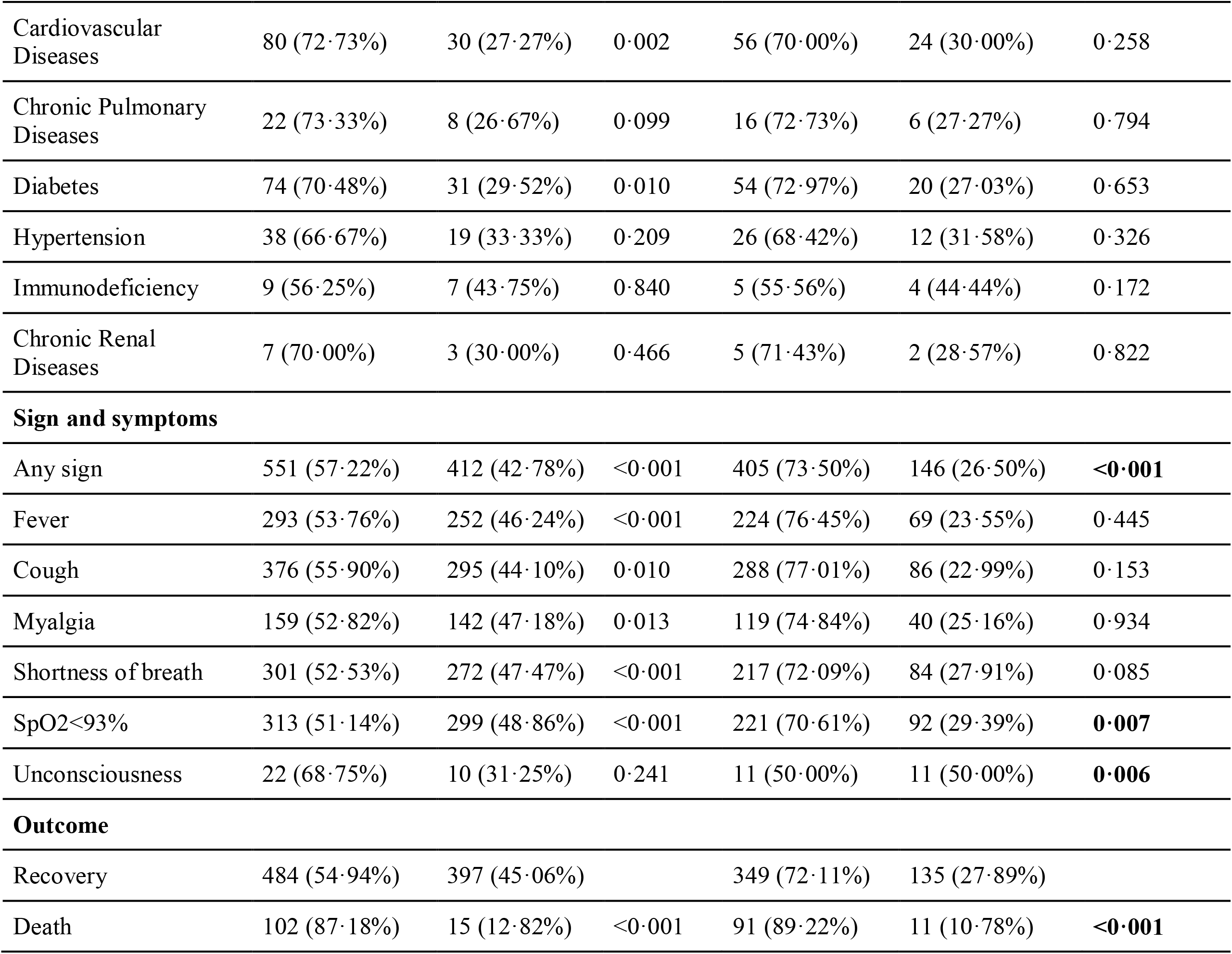
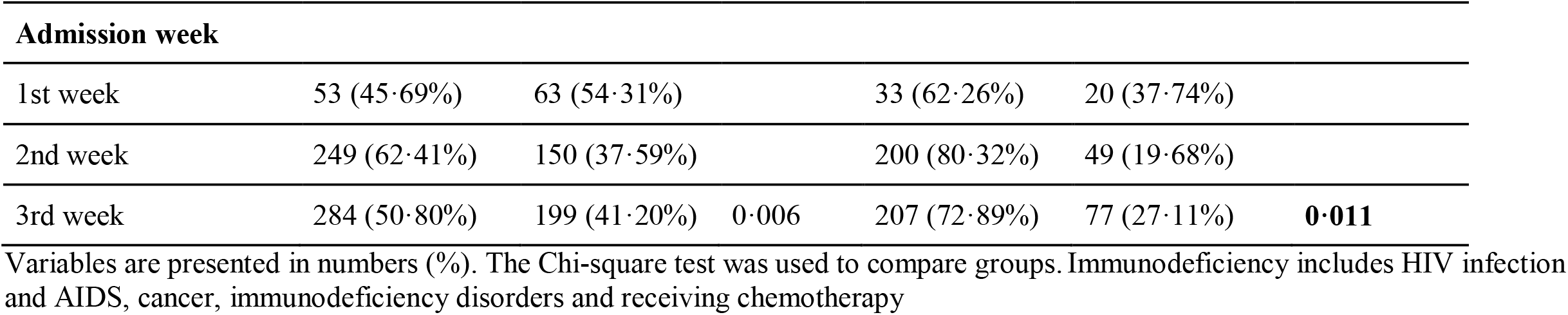
Comparing the study patients based on their PCR-test status

The case fatality rate among admitted patients in Qazvin province was 11·7% (Table 1). However, the number differed between various groups (Table 4). Employing a multiple logistic regression model including age group, sex, type of hospital (teaching, non-teaching (high volume and low volume), week of admission, ICU admission, co-existing disorders and rRT-PCR test results to determine the associated factors of death in the patients showed that age>70 years (OR=5·2 95% CI 2·9-9·1, p<0·001), ICU admission (OR=11·5, 95% CI 5·5-23·9, p<0·001) and having positive rRT-PCR test results (OR=5·8, 95%CI 2·7-12·5, p<0·001) were the main determinants of death in the patients. After adjustment for all confounding factors including sex, age, ICU admission, co-exiting disorders, and weeks of admission, higher odds of death was observed in rRT-PCR positive cases compared to test-negative patients (OR=5·7, 95% CI 2·6-12·4, p<0·001) (Table 4).

**Table 4:** Associated factors of death in COVID-19 patients who were admitted to hospitals in Qazvin province

| Subgroup | Number of death (%) | Odds ratio (95% CI) | P value |
| --- | --- | --- | --- |
| **Sex** |  |  |  |
| Female | 48 (11·19%) | Reference |  |
| Male | 69 (12·13%) | 1·16 (0·74, 1·81) | 0·511 |
| **Age group** |  |  |  |
| < 50 years | 24 (5·97%) | Reference |  |
| 50-59 years | 12 (6·25%) | 0·97 (0·44, 2·11) | 0·942 |
| 60-69 years | 22 (11·76%) | 1·73 (0·89, 3·35) | 0·101 |
| ≥70 years | 59 (27·19%) | 5·15 (2·91, 9·12) | **<0·001** |
| **ICU admission** |  |  |  |
| No | 92 (9·73%) | Reference |  |
| Yes | 25 (48·08%) | 11·46 (5·53, 23·86) | **<0·001** |
| **RT-PCR test status** |  |  |  |
| Negative | 11 (7·53%) | Reference |  |
| Positive | 91 (20·68%) | 5·75 (2·65, 12·48) | **<0·001** |
| Not tested | 15 (3·64%) | 0·74 (0·30, 1·84) | 0·531 |
| **Co-existing disorder** |  |  |  |
| Without co-existing disorder | 67 (9·14%) | Reference |  |
| All | 50 (18·87%) | 1·4 (0·91, 2·34) | 0·110 |
| Two or more co-disorders | 20 (25·02%) | 1·82 (0·92, 3·60) | 0·081 |
| Cardiovascular diseases | 26 (23·64%) | 1·57 (0·86, 2·87) | 0·186 |
| Chronic pulmonary diseases | 5 (16·67%) | 1·28 (0·43, 3·79) | 0·438 |
| Diabetes | 19 (18·10%) | 1·39 (0·73, 2·64) | 0·287 |
| Chronic renal disease | 4 (40·00%) | 3·07 (0·55, 17·08) | 0·164 |
| Immunodeficiency | **5 (31·25%)** | **4·3 (1·11, 17·23)** | **0·035** |
| Hypertension | 12 (21·05%) | 0·99 (0·44, 2·20) | 0·991 |
| **Admission week** |  |  |  |
| 1<sup>st</sup> Week | 12 (10·34%) | Reference |  |
| 2<sup>nd</sup> Week | 47 (11·78%) | 0·77 (0·32, 1·75) | 0·536 |
| 3<sup>rd</sup> Week | 58 (12·01%) | 0·92 (0·41, 2·05) | 0·845 |
| **Type of Hospital** |  |  |  |
| Teaching Hospitals | 93 (12·09%) | Reference |  |
| High-Volume non-teaching | 22 (10·38%) | 0·61 (0·32, 1·11) | 0·140 |
| Low-Volume non-teaching | 2 (11·76%) | 0·23 (0·03, 1·55) | 0·113 |

Multivariate logistic regression analysis was run to calculate odd ratios (95%CI). Immunodeficiency includes HIV infection and AIDS, cancer, immunodeficiency disorders, and receiving chemotherapy; Low volume hospitals indicate hospitals with less than 20 COVID-19 admitted patients.

## Discussion

The present study which was conducted in the critical first 3 weeks of the novel coronavirus outbreak in Qazvin province, Iran, found that while CT-diagnosed COVID-19 patients with typical respiratory symptoms but negative rRT-PCR results had a 7·5% case fatality rate, the number was as high as 20·7% in the test-positive patients.

This study showed a similar case fatality rate between men and women (12·13% vs 11·19%, p=0·511). This is in line with what has been previously reported from Wuhan, China.^(4)^ On the other hand, as was previously presumed, the case fatality rate in patients aged > 70 years (OR=5·2, 95%CI 2·9-9·1) and in those who were ICU-admitted (OR=11·5, 95%CI 5·5-23·9) were higher when compared to the reference group. Nevertheless, we did not find any statistically significant increased death rate among COVID-19 patients with co-existing diseases. The exception, however, was those with immunodeficiency disorders whose case fatality rate was about 4 times the reference group. (Table 3) In fact, in the regression model, age affected death both independently and through the “corridor” of comorbidities. On the other hand, the adverse effect of major comorbidities was apparently observed only in association with age, i.e., they had no independent role in the case fatality rate in this cohort of patients. On the contrary, Guan et al previously reported that among PCR-test-confirmed hospitalized patients, the composite endpoint including admission to ICU, invasive ventilation, and death was higher in those who had known comorbidities.^(5)^ Also, one meta-analysis previously reported that certain comorbidities were more prevalent in severe COVID-19 patients.^(6)^

This study is among the first investigations that aimed to frame the current picture of the battle with the COVID-19 outbreak in one of Iran’s provinces in the first 3 weeks of the epidemic emergence. We consider the timing of this study - reporting the clinical pattern of hospitalized patients in an early period when the medical system is assumed not to have become overloaded yet - to be among the strengths of this study. Additionally, acquiring data from the electronic data entry platform, which was designated exclusively to collect the data of the COVID-19 patients during their course of admission, along with the sizable number of the patients, add to the robustness of this study results. However, the findings on the clinical pattern and outcome of the patients need to be interpreted in the context of our limitations. Among them, we need to emphasize on the low availability of PCR testing in Iran. Also we did not have access to data on the patients’ laboratory results, their smoking status, or the medications they received.

The applied diagnosis and treatment flowchart as was first proposed by the INIGCDT^(3)^ merits further explanation. The guidance which had originally adapted the related World Health Organization (WHO) guidance,^(7, 8)^ proposed chest imaging as the first diagnostic step to screen patients who require prompt hospitalization amid shortages in RT-PCR test kits in Iran. As a result, all hospital-admitted patients with suggestive SARS-CoV-2 symptoms - with or without comorbidities - underwent chest imaging (specifically chest CT-scan) in the province of Qazvin hospitals. From among them, 59·3% were tested with RT-PCR and 24·9% had negative results. The proportion of the negative test results is in line with previous findings. While there has been a significant correlation between throat swab and sputum sample viral loads,^(9)^ one study examining the bio-distribution of SARS-CoV-2 in different body tissues, reported positive RT-PCR rates only in 72% of sputum specimens.^(10)^

Examining the concordance between chest CT-scan and PCR test results, a previous study from Wuhan, China, reported that chest CT sensitivity was 97% in RT-PCR-positive patients. On the other hand, in the PCR-negative patients, 75% had positive CT scan findings, 81% of whom were later considered as highly likely or probable cases in their study. ^(11)^ Another study also showed that the sensitivity of chest CT was significantly higher than RT-PCR in their patients (98% vs 71%).^(12)^

When categorizing the CT-diagnosed COVID-19 patients based on their RT-PCR test results, as expected, the worst scenario was reported in the group of patients with suggestive respiratory symptoms or underlying diseases who had positive PCR test results. They accounted for 41% of ICU admissions throughout the province with a 77% case fatality rate even after receiving ICU care. On the other hand, the group with negative PCR test results who had presented with suggestive clinical symptoms or had had underlying diseases accounted for 32% of ICU admissions due to COVID-19 in the province, among them 71% recovered. It has been previously reported that negative PCR results might be due to lower viral load in patients’ specimens.^(13, 14)^ This might provide a possible explanation for the lower case fatality rate among the test-negative group.

In the “no-test” group, which comprised CT-positive patients who were not tested for RT-PCR despite having suggestive clinical symptoms or co-existent diseases, 3·6% were admitted to intensive care units, among which 66·7% recovered eventually. On the other hand, the overall case fatality rate between this no-test group and those with negative PCR results did not differ significantly (4·1% vs 7·5%). Considering that in the first weeks of the epidemic, PCR testing was reserved for patients in critical condition, one might presume that a rather preemptive transfer of patients to the ICU and providing them with early intensive care would partially explain the lower case fatality rate in this group.

Additionally, there was a group of study patients with negative PCR results who also lacked major clinical symptoms or significant comorbidities. These patients had the lowest case fatality rate (3·1%) and none of them were ICU-admitted. These non-COVID-19 hospitalized patients who were not included in the analysis may serve as a basis for comparing the admission course and outcomes of the study patients.

The importance of intensive care facilities availability in such an epidemic cannot be emphasized more. As officially announced by the Iran Ministry of Health, in 2018, there were a total of 8264 ICU beds nationwide, which were occupied by 453891 patients during the year-ending 2018 (occupancy rate 55 patients/bed/year). In Qazvin province, which includes 1·6% of Iran’s population, there are 96 ICU beds available (7·7 beds/100,000 of the population), comprising 1·1% of the total ICU beds in Iran. In this study, among the 92 non-survivor individuals that were treated outside intensive care units, 63% presented with shortness of breath and 66% had low pulse oximetry readings. On the other hand, among the 612 patients who presented with SpO2 < 93%, only 6·2% were admitted to intensive care units. Considering that the ICU admission rate in this study was 5·2% - i.e., 52 ICU beds out of the Qazvin province total ICU beds, it could be concluded that there has been a possible need for at least 90 additional ICU beds throughout the province, that if had been provided, more lives would be saved. Interestingly, other studies have previously reported that while most people affected by COVID-19 present with a mild illness, about 14% progress to more severe forms of the disease that require hospitalization, and 5% need ICU care.^(15-17)^

We also need to briefly mention the process of RT-PCR testing in the study patients. Due to the considerable shortage in PCR test kits in Iran and according to the INIGCDT, physicians were encouraged to perform the test only in patients with critical conditions. Moreover, no re-testing was provided for patients with negative tests. While it has been reported that COVID-19 patients with more severe forms of the disease have higher viral loads and longer periods of viral-shedding,^(7)^ testing protocol mentioned above might have affected the results on the association between positive PCR test results and case fatality rate as well.

In conclusion, this study observed that COVID-19 hospitalized patients with mild symptoms despite positive chest CT changes and major comorbidities were more probable to have negative rRT-PCR test results, hence a lower case fatality rate and a more favorable outcome. Conversely, positive rRT-PCR test results were more prevalent in patients presenting with low SpO2 or unconsciousness and was strongly associated with increased odds of death among chest CT-positive patients. Considering the serious shortage in ICU capacity, preemptive transfer of more vulnerable rRT-test-positive patients to the ICU might save their lives.

## Data Availability

All data referred to in this manuscript is available upon request.

## Manuscript preparation

WE4H writing and editing services, Vancouver, Canada, provided medical writing and editing support in preparing this manuscript.

